# Understanding the current and future usage of donor human milk in hospitals: an online survey of UK neonatal units

**DOI:** 10.1101/2022.12.23.22283892

**Authors:** NS Shenker, S Griffin, J Hamill-Keays, M Thomson, J Simpson, G Weaver

## Abstract

**Objective:** The use of donor human milk (DHM) where there is a shortfall of maternal milk can benefit both infant and maternal outcomes but DHM supply is not always assured. This study aimed to understand current DHM usage in UK neonatal units and potential future demand to inform service planning.

**Design/Setting:** An online survey was disseminated to all UK neonatal units using SmartSurvey or by telephone between February and April 2022 after development alongside neonatal unit teams.

**Results:** Surveys were completed by 55.4% units (108/195; 18 Level 1, 47 Level 2, 41 Level 3; cot numbers 9-56) from all thirteen Operational Delivery Networks. Only four units reported not using DHM, and another two units only if infants are transferred on DHM feeds. There was marked diversity in DHM implementation and usage. Five of six units with their own milk bank had needed to source milk from an external milk bank in the last year. Ninety units (84.9%) considered DHM was sometimes (n=35) or always (n=55) supportive of maternal breastfeeding, and rarely supportive by three (2.9%). Usage was predicted to increase by 37 units (34.9%), driven by parental preference, clinical trials, and improved evidence.

**Conclusions:** These findings support the assumption that UK hospital DHM demand will increase after updated recommendations from the WHO and British Association of Perinatal Medicine. This data will help policymakers and milk banks to plan strategic service delivery, alongside ongoing cost-benefit analyses, donor recruitment strategies and infrastructure planning to ensure equity of assured access to DHM nationally.

**What’s known, what does this study add?:**

- **What is already known on this topic** Updated recommendations from WHO and BAPM are likely to increase demand for donor human milk provision from human milk banks, but there have been no recent data collected on baseline usage criteria, enteral feeding guidelines, and anticipated future use.
- **What this study adds** This national survey of UK neonatal units highlights variability in DHM provision, and reasons for demand spikes that will be helpful for modelling future services. Almost 85% of neonatal units responded that DHM availability was supportive of maternal lactation support. DHM demand is likely to increase further in the next 2 years.
- **How this study might affect research, practice or policy** Understanding demand variability will help in planning nationally equitable human milk bank services, and support the development of robust national service continuity planning. The study also highlights variability in practice, often in single regions, raising concerns related to health equity, staffing limitations and uncertainty in DHM implementation.

## Introduction

Donor human milk (DHM) describes milk that is collected from screened donors, heat-treated and screened microbiologically before being dispatched to end users in hospitals or in the community. DHM is used as a temporary bridge to breastfeeding, usually for extremely vulnerable premature infants at high risk of necrotising enterocolitis where early feeds are preferable but there is a shortfall in maternal milk supply (1, 2). Recent *“World Health Organization (WHO) recommendations on the care of preterm or low birth-weight infants”* recommend with moderate certainty that DHM should be available for all preterm infants where there is insufficient maternal milk (3). When UK DHM use was last examined, a report by the British Association of Perinatal Medicine (BAPM) on the provision of human milk banking (HMB) services in the UK in 2016 recommended that more research would be needed before national recommendations on the planning and funding of nationally equitable human HMB services could be made (4). Since then, new avenues of research have established that, when used in the context of optimal lactation support, DHM can be supportive of the establishment of maternal lactation, improved parental wellbeing, maternal psychological health and infant outcomes (5, 6, 7, 8, 9). DHM availability may also have positive impacts on infant and maternal health beyond the preterm population (7, 10, 11, 12).

During 2022 a BAPM Framework for Practice working group developed new recommendations for the use of DHM within UK hospitals (13). Research continues into the efficacy of DHM availability alongside lactation support for the above outcomes in postnatal wards, late and moderately preterm infants, those with other health issues, and full-term healthy infants where mothers face developmental or other challenges in establishing a full milk supply, or where the choice to breastfeed is not possible.

DHM demand from neonatal units can fluctuate markedly, leaving NHS milk banks occasionally unable to meet the demand. Increasing uncertainties and risks in recent years, including the COVID-19 pandemic, fuel shortages, and cost of living increases, have made the operation of smaller services difficult both in the UK and globally (13, 14). Indeed, national reductions in milk donor numbers and increased demand from hospital NICUs in the final quarter of 2021 led to six NHS milk banks running out of DHM stock (personal correspondence, UKAMB 2022), including large milk banks that supply regionally. As well as understanding drivers of demand spikes, gaps in knowledge exist on individual neonatal unit practices including DHM guideline use, eligibility criteria, and how neonatal teams predict the use of DHM will change in the future. This online mixed-methods survey was designed to investigate causes of demand fluctuation, current guidelines for DHM and the likelihood of DHM demand increasing or decreasing in future. This study, the first national survey of DHM use since 2015 (15), aimed to assist future UK HMB service planning.

## Methods

A survey was developed with joint expertise from neonatologists (MT and JS), milk banking experts (GW, JHK) and academics (NS), and piloted by eight neonatal units by phone. The survey was then disseminated by email to all 195 UK Neonatal Units online via an Imperial College London Qualtrics license (Qualtrics^XM^, Seattle, WA, USA). Three follow-up email reminders were sent and phone reminders where possible in the study period from February 2022 to May 2022.

Data was exported into Excel (Microsoft Excel, Washington, USA). Descriptive statistics were calculated as percentages and statements regarding the future anticipated use of DHM on neonatal units were collated and categorised thematically.

## Results

### Responses

Of the 195 neonatal units, responses were submitted from 137 units (70.2%) with full responses suitable for analysis returned for 108 units (55.4%). Two responses included data for more than one unit, and for the purposes of analysis these were treated as a single unit (n=106). Of these, 18 (17.0%, 211 cots) were Level 1 units, 47 Level 2 (44.3%, 920 cots) and 41 Level 3 units (38.7%, 1313 cots). There were regional disparities in response rates, from 27.8% of all units responding from the Yorkshire and Humber Operational Delivery Network (ODN) to 100% of units responding in the Southwest ODN (Supp Table 1). Responses were completed by a range of healthcare professionals, reflecting the range of input into infant feeding decisions and responsibility in different units, with 74% of respondents identifying as having responsibility for the infant feeding guideline in their hospital (Table 1).

**Table 1:** Healthcare professional completing the survey, with or without specific responsibility for infant enteral feeding.

|  | **No. responses (%)** | **Specific infant feeding responsibility**<br>**(% of responses)** |
| --- | --- | --- |
| Neonatal Nurse | 31 (29.2) | 21 (67.7) |
| Infant Feeding Lead | 20 (18.9) | 20 (100) |
| Ward Manager | 20 (18.9) | 12 (60) |
| Neonatal Dietitian | 15 (14.2) | 14 (93.3) |
| Consultant Neonatologist | 14 (13.2) | 9 (64.3) |
| Consultant Paediatrician | 5 (4.7) | 1 (20.0) |
| Paediatric registrar | 1 (0.9) | 1 (100) |
| **Total** | **106** | **78** |

### Infant feeding leads and guidelines

Forty-five units reported having a dedicated infant feeding specialist (infant feeding lead, IFL) on their unit (42.9%), 35 (33.3%) had an IFL whose workload was shared with maternity and/or paediatric services, 17 (16.2%) had no IFL but reported maternity and/or paediatrics departments did, and 8 (7.6%) had no IFL within the Trust. Overall, 102 (96.2) units had an enteral feeding guideline, of which 72 (67.9%) units had updated within the last 3 years. Fifty-six (58.9%) units had a specific DHM use guideline (45 updated within the last 3 yr), and 71 (67%) units had a specialist neonatal dietician. Most guidelines were adapted from the unit’s ODN guidelines (n=60, 56.6%), with a further 33 units (31.1%) use a guideline written by individual Trust staff. Thirteen responses (12.3%) were unsure how their Trust guideline had been developed.

### Use of donor human milk (DHM)

Of the 106 units that responded, 97 (91.5%) used DHM. Five (4.7%) reported only using DHM if an infant was transferred while being fed with DHM, and four (3.8%) did not use DHM at all. Of the indications for DHM within unit enteral feeding guidelines, the commonest were infants under a specified gestational age (87/102, 85.3%), use for infants under a specific birth weight (77/102, 75.5%; Table 2).

**Table 2:** Indications for DHM within unit enteral feeding guidelines, from 102 neonatal unit responses.

| Enteral feeding guideline indication for DHM | No. units including indication in enteral feeding guideline (%) |
| --- | --- |
| Under a specified gestational age (weeks) | 87 (85.3) |
| Under a specific birth weight (g) | 77 (75.5) |
| Reversed or persistently absent end diastolic flow | 56 (54.9) |
| Post-medical NEC | 53 (52.0) |
| Post-surgical NEC | 52 (51.0) |
| Parental preference | 23 (22.5) |
| Congenital bowel anomaly e.g., gastroschisis | 23 (22.5) |
| Cardiac anomaly | 23 (22.5) |
| Top up (bridging) feeds | 22 (21.6) |
| Haemodynamically unstable / inotropic support | 19 (18.6) |
| HIE | 17 (16.7) |
| PDA | 12 (11.8) |
| Parental allergies | 2 (2.9) |
| Other* | 40 (39.2) |

NEC: Necrotising enterocolitis; HIE: hypoxic-ischaemic encephalopathy; PDA: patent ductus arteriosus.

Forty units used DHM for additional indications, including to support maternal efforts to establish breastfeeding, parental preference, maternal HIV, bilateral mastectomy, if a twin, triplet or quadruplet was receiving DHM for another indication, maternal medication contraindicated to breastfeeding, and consultant discretion. One unit reported using DHM outside of the protocol if the use-by-date of the DHM was approaching. Five units reported using DHM in the context of a research trial. Several respondents reported that parental preference was not included in the current guidance but would be included in the next.

When asked whether the availability of DHM was supportive of lactation and breastfeeding on their neonatal unit, 55 units (53.9%) responded it was always supportive, 35 units (34.3%) felt it was sometimes supportive, eight units (7.8%) were unsure and three units (2.9%) reported it frequently undermined maternal breastfeeding. Overall 90 units (88.2%) felt DHM was supportive of breastfeeding.

In the last year, 88 units had sourced DHM from an external milk bank, six only used DHM from a milk bank within their Trust. Of these, five had needed to source milk externally because of a lack of supply from their milk bank (information about individual milk bank provision in Supplemental Table 3).

Participants were asked to estimate the requirement for DHM over the next 2 years. Thirty-seven units (34.9%) felt that their requirement would increase, 33 units (31.3%) predicted usage would stay the same, and four units (3.8%) reported usage would decrease. Twenty-eight responses were unsure. Factors that underpinned these responses are reported in Table 3, with the lead cause for DHM requirement increasing being a change in the criteria for patients eligible for DHM, followed by increasing admissions and parental request. The major cause of decreased use would be reduced availability of DHM, and increased breastfeeding rates. Additional written explanations described increased staff awareness in the role of DHM in supporting the health of premature infants and greater parental awareness of the benefits of both maternal milk provision and DHM supplementation. Respondents concerns commonly included lack of assured supply, milk banks running out, and the need for neonatal infant feeding leads to embed systems, guidelines and unit cultural change.

**Table 3:** Factors contributing to changed DHM use over the next 2 years.

|  | Increasing | Staying the same | Decreasing | NA | No response |
| --- | --- | --- | --- | --- | --- |
| No. patients | 19 | 58 | 1 | 20 | 8 |
| Changing mix of patients | 14 | 52 | 4 | 26 | 10 |
| Criteria for patients eligible for donor milk changing | 29 | 48 | 4 | 17 | 8 |
| Number of parents or carers requesting donor milk | 17 | 53 | 1 | 28 | 7 |
| Availability of donor milk | 16 | 55 | 10 | 17 | 8 |
| Change in funding for donor milk | 1 | 66 | 1 | 29 | 9 |
| Involvement in clinical trials or quality improvement studies | 9 | 31 | 0 | 50 | 16 |

## Discussion

This study was an online mixed-methods survey of DHM usage on neonatal units in the UK. The major findings are that DHM is used in the majority of units that responded, increased from those reported previously (15, 16). Most units anticipated continuing to use DHM at current or increasing volumes, and that DHM requirements would increase as rationing reduced, contingent on availability. The survey highlights demand spikes and supply issues related to a range of issues, including increased birth rates, births of multiples, engagement by neonatal units in clinical trials requiring additional DHM, and broadening of DHM use criteria.

While not the primary purpose of the survey, the results highlight widespread variability in DHM use, eligibility criteria, guidelines and staffing to support appropriate use in the context of optimal lactation support. These results are consistent with the Neonatology GIRFT Programme National Specialty Report (17), which reported differences in access to DMH between Trusts in the same ODN, and marked variability between ODNs. As supported in this survey, only one ODN does not support the use of DHM at all.

Operationally, this survey highlights some of the needs of NNUs that could be reflected in optimal milk bank service provision. Ensuring an adequate use by date would reduce wastage from discarded DHM, and whilst some milk banks guarantee a minimum use-by-date of 8 weeks, this is not widespread practice. Nine responses reported that DHM use would increase as further clinical trials were implemented, and five responded that the FEED1 clinical trial, examining the impact of early feeding on infant outcomes (18), had increased DHM usage on their unit. Early and comprehensive communication and planning between trial teams and milk banks is necessary to ensure intervention provision can be guaranteed. The response from five units that DHM was not always available from their own Trust or regional milk bank highlights the need for strengthened services. HMB services need to be able to guarantee assured provision to meet increased demand from national and global recommendations, ensuring clinicians can confidently advise parents and caregivers. To ensure service resilience, appropriate consideration is also needed of service-specific risks and the design and implementation of a HMB-specific risk register and full service continuity plans (19).

The majority of respondents (88.2%) reported that they were confident that the use of DHM was supportive of lactation and breastfeeding on their NNU. One of the key arguments against the use of DHM has been that its availability could reduce the motivation of mothers to establish their own milk supply (20). Randomised control trials in India (5), along with observational findings from the USA (6, 8, 9), India (21) and Europe (7, 22), published since this systematic review have confirmed that when used in the context of optimal lactation support, DHM can be additionally supportive of lactation. This consistent finding could be explained in part by the psychological stressor of trying to avoid formula use and with the widely known risks of increased NEC and other complications of prematurity, which may impact negatively on the physiological ability of mothers to establish lactation (7, 23, 24). Secondary lactogenesis depends on an increase in prolactin and oxytocin, mediated by the hypothalamic-pituitary axis. The stress hormone cortisol is a negative inhibitor of prolactin release (25), and amongst the stress of preterm birth and NICU stay, the additional stressor of being responsible for producing a full feed requirement immediately, particularly if the mother is unwell herself, may undermine maternal milk production. A limited qualitative study from the US with 35 participants highlighted that women felt DHM offered a bridge to breastfeeding, allowing time to overcome short term hurdles, while the introduction of infant formula compounded their feelings of personal failure (26). In a recent sample of 105 parents whose infants have received DHM within the last year, responses suggest increased parental wellbeing (7), though further studies are needed to understand how widely these perceptions may be in the general population.

The study strengths included a high response rate, particularly for Level 2 and 3 NNUs, but also a good geographical spread that was representative of each neonatal ODN. The study also included responses from a high proportion of healthcare professionals with responsibility for infant feeding on the NNU (n=78, 74%), giving confidence in the accuracy of the data collected. This study was limited by the nature of the design as an online questionnaire, meaning more work is needed in interviews to understand the barriers and motivations behind the increased use of DHM on neonatal units.

In conclusion, demand for DHM on hospital neonatal units is increasing, and predicted to increase further in the majority of units over the next 2 years. Indications vary markedly between units, and nationally agreed recommendations are urgently needed to reduce inequities in access to DHM. This study supports the findings from previous studies that have established the impact on breastfeeding rates on discharge of combined lactation support and DHM availability, as well as overall parental wellbeing, with over 88% of responses reporting that DHM availability was supportive of breastfeeding. Enhanced lactation interventions and parental wishes are likely to play a key role in the increased demand, and use of DHM should decrease as breastfeeding support increases, meaning broadened use criteria could be considered. Further work is needed to understand demand estimation, optimal models for donor milk provision, and clinical trials to determine efficacy of use in a wide range of clinical scenarios. Acceptability and scalability assessments, as well as health economic studies into cost-effectiveness and service costing, are currently underway and aim to provide data to inform funding of future nationally equitable milk bank services within the next 2 years.

## Data Availability

All data produced in the present study are available upon reasonable request to the authors.

## Acknowledgements

The authors are grateful to the healthcare professionals in neonatal units across the UK who took the time to develop and complete this questionnaire.

## Authorship contribution

NS and GW conceived the study. NS, GW and MT developed the questionnaire, and JS approved contents. JHK and NS conducted data collection. NS conducted data analysis. All authors contributed to manuscript writing and interpretation of results. All authors approved the completed manuscript before submission.

## Supplemental Information

**Supplemental Table 1:** Survey response rates from across each of the Operational Delivery Networks in the UK.

| ODN | Total responses | Level 1 | Level 2 | Level 3 | No. units in ODN | Response (%) |
| --- | --- | --- | --- | --- | --- | --- |
| East Midlands | 4 | 0 | 2 | 2 | 11 | 36.4% |
| East of England | 14 | 3 | 6 | 5 | 17 | 82.4% |
| KSS | 7 | 3 | 2 | 2 | 16 | 43.8% |
| London | 16 | 1 | 11 | 4 | 27 | 59.3% |
| North West | 7 | 2 | 1 | 4 | 22 | 31.8% |
| Northern | 5 | 1 | 0 | 4 | 10 | 50.0% |
| Northern Ireland | 3 | 2 | 0 | 1 | 7 | 42.9% |
| Scotland | 5 | 0 | 2 | 3 | 15 | 33.3% |
| South West | 12 | 2 | 6 | 4 | 12 | 100.0% |
| Thames Valley and Wessex | 10 | 2 | 6 | 2 | 14 | 71.4% |
| Wales | 7 | 0 | 3 | 3 | 12 | 58.3% |
| West Midlands | 13 | 2 | 6 | 4 | 14 | 92.9% |
| Yorkshire and Humber | 5 | 0 | 2 | 3 | 18 | 27.8% |
| **Total** | **108** | **18** | **47** | **41** | **195** | **55.4%** |

**Supplemental Table 2:** Indications for the use of DHM included in enteral infant feeding protocol.

| **Indications** | **Yes** | **No** |
| --- | --- | --- |
| Under a specified gestational age (weeks) | 87 | 15 |
| Under a specific birth weight (g) | 77 | 25 |
| Reversed or persistently absent end diastolic flow | 56 | 46 |
| Post-medical NEC | 53 | 49 |
| Post-surgical NEC | 52 | 50 |
| Parental preference | 23 | 79 |
| Congenital bowel anomaly e.g., gastroschisis | 23 | 79 |
| Cardiac anomaly | 23 | 79 |
| Top-up (bridging) feeds | 22 | 80 |
| Haemodynamically unstable / inotropic support | 19 | 83 |
| HIE | 17 | 85 |
| PDA | 12 | 90 |
| Parental allergies | 2 | 100 |
| **Other** | **40** | **62** |

\* Additional indications included supporting maternal efforts to establish breastfeeding, parental preference, maternal HIV, bilateral mastectomy, if a twin / triplet / quad was receiving DHM for another indication, maternal medication contraindicated to breastfeeding and consultant discretion. Several reported that parental preference was not included in the current guidance but would be included in the next.

**Supplemental Table 3:** Reported hospital usage of UK milk banks from the survey responses of hospitals that do not have a milk bank in their own hospital.

| **Milk bank** | **Number of units hospitals supplied by external milk banks\*** |
| --- | --- |
| Hearts | 27 |
| Birmingham | 12 |
| Chester | 11 |
| Southampton | 7 |
| Oxford | 6 |
| Cambridge | 5 |
| Calderdale | 2 |
| STTH | 1 |
| St Georges | 1 |
| QCCH | 1 |
| **Total** | **86** |

\*In addition to individual Trust milk banks which may supply hospitals within their ODN and beyond, the Scottish National Service provides equitable access to each of the 15 NNUs in all Scottish Health Boards, the South West Acute Hospital Human Milk Bank in Northern Ireland is the first line milk bank for all NNUs in Northern Ireland (n=7) and the Republic of Ireland (n=19), and the Southwest Human Milk Bank in Bristol is funded to supply all units in the southwest ODN (n = 12).

